# Prism Adaptation-Induced Modulation of Cortical Excitability of Upper and Lower Limb Muscles is Enhanced with Electrical Stimulation

**DOI:** 10.1101/2024.09.30.24314639

**Authors:** Fisayo K Aloba, Jasmine M Hope, Jacob Spencer, Maithri Muthukumar, Taylor M Leone, Vyoma Parikh, Peii Chen, Michael R Borich, Trisha M Kesar

## Abstract

**Background:** Prism adaptation (PA) is a sensorimotor behavioral phenomenon. Right shifting PA induces a shift in global visuospatial motor behavior toward the left hemi-space (aftereffect) leading to immediate and transient changes in visuomotor behavior. Non-invasive sensorimotor electrical stimulation (Stim) may upregulate corticomotor excitability, is commonly used as a therapeutic adjunct during motor training, and may accentuate the effects of PA. However, the cortical plasticity mechanisms related to the behavioral effects of PA, its generalization to the lower limb, and the combinatorial effects of PA and Stim are poorly understood.

**Objective:** To evaluate the effects of combining PA with Stim on corticomotor excitability and visuo-spatial-motor behavior, and its generalization to the lower limb.

**Methods:** We used a repeated-measures design to evaluate the effects of 1 session of PA with and without Stim in 15 young able-bodied individuals (18-35 years). Before and after PA, visuomotor pointing task performance, corticomotor excitability, short-interval intra-cortical inhibition (SICI), long-interval intra-cortical inhibition (LICI), and intra-cortical facilitation (ICF) were evaluated in bilateral upper and left ankle muscles with motor-evoked potentials (MEPs) elicited from single and paired-pulse transcranial magnetic stimulation (TMS).

**Results:** Behaviorally, both PA+Stim and PA+Sham showed significant sensorimotor aftereffects, inducing a leftward shift in visuo-spatial and proprioceptive pointing. Neurophysiologically, suprathreshold MEP amplitude increased in the left first dorsal interossei (FDI) and left soleus following the PA+Stim condition but not the PA+Sham condition. PA+Stim showed statistical trends for inducing larger changes in ICF of the left FDI and left tibialis anterior. Additionally, compared to PA+Stim, PA+Sham induced larger changes in LICI of the left FDI and left tibialis anterior, and in SICI of the left tibialis anterior.

**Conclusion:** Although both PA+Stim and PA+Sham had similar behavioral aftereffects, only PA+Stim increased cortical excitability in M1 representations of the left upper and lower limb (toward the direction of the PA aftereffect), suggesting that PA+Stim may elicit greater neurophysiological changes and generalization to lower limb than PA alone.

## 1. Introduction

Visuospatial-motor deficits are complex disorders involving the visual, perceptual, sensory, and motor systems^1,2^. These deficits are caused by dysfunctions in visuospatial and sensorimotor circuits of the brain that integrate visual and sensory inputs with motor commands to enable motor skills needed to carry out simple functional tasks such as reaching for a coffee cup or walking^1^. Prism adaptation (PA) is a sensorimotor paradigm used as a behavioral task to assess visuomotor adaptation capacity. Multiple sessions of PA are also used as tool to modulate higher spatial cognitive processes^6,7^ and improve visuo-motor function. Right-shifting PA involves wearing prism lenses that shift visuospatial behavior towards the right hemispace, and the aftereffect is a perceptual and motor bias toward the left hemispace. The PA protocol consists of three phases. Before prisms are put on, the participant points at visual targets to obtain reference values (pre-adaptation pointing performance). When wearing prism lenses (*prism exposure)* that induce a rightward visual displacement of the entire visual field, during early adaptation, the participant initially incorrectly points to the right side of the target (because the image of the target has shifted to the right). The visual feedback of the error is immediately available to the participant. To correct this “error”, the participant points leftward until perceived correct pointing to the object, which occurs by the late adaptation period. When prism glasses are removed, due to the sensorimotor adaptation, the participant continues aiming towards the left side of visual targets— measured as the prism aftereffect.^6,8–13^ These effects of PA on visuo-motor behavior have been investigated in healthy individuals and also applied to individuals with spatial neglect.^6,14–16^ Studies have shown that 1-2 sessions of PA can induce changes in visuo-spatial-motor behavior, with effects lasting from 24 hours to one week^6,16–18^ These not only include improved perceived body-midline but generalize to other activities such as wheelchair navigation and transfers^17,19–22^

Visuo-spatial cognitive information processing involves the input stage, which comprises perception and attention, the representational stage comprising sensorimotor integration, and the motor output stage involving intentional movements. Different theories about how PA affects the various stages of cognitive information are debated. Adapting to right-shifting prisms leads to visuospatial motor behavior toward the left hemi-space after prism removal. Previous literature suggests that this aftereffect is induced through changes in the dorsal fronto-parietal networks (superior parietal lobule, intraparietal sulcus, and dorsolateral prefrontal cortex, which are responsible for spatial processing and attention control, and are connected to the primary motor cortex^48^. PA is thought to recalibrate spatial representation and strengthen the connection between perception and motor actions thereby influencing both lower-level sensorimotor behavior and higher cognitive processes like spatial awareness and attention^6,23^. However, other studies suggest that PA may primarily affect the motor-output stage^16,24,25^. The lack of clarity regarding the neural mechanisms underlying PA and high inter-individual variability in the magnitude of PA-induced behavioral effects calls for more investigation to determine the optimal protocol and neural mechanisms of PA.

Somatosensory electrical stimulation is a promising and accessible intervention to improve spatial representation and enhance the somatosensory inputs from the neglected or paretic side of the body^26^ in post-stroke spatial neglect^27–29^. Somatosensory stimulation may modulate the transformation of spatial coordinates needed for visuo-spatial orientation^26^. Previous studies have used different stimulation techniques such as neck vibration^30^ and transcranial direct current stimulation^31^ (tDCS) in combination with PA to improve visuospatial and sensory deficits. Previous research shows that sensory stimulation delivered to a dermatome can increase the excitability of the primary motor cortex (M1) of the corresponding myotome ^32–36^. Similarly, excitatory tDCS combined with PA modulates upper limb M1 excitability in young healthy adults^37^. Thus, sensory stimulation can prove to be a valuable adjunct to PA, helping improve the consistency and magnitude of the effects of PA on visuospatial behavior.

Understanding the neural mechanisms of how paradigms such as PA induce behavioral change is a necessary prerequisite to designing more personalized and effective rehabilitation treatments. Transcranial magnetic stimulation (TMS) is a non-invasive tool commonly used to evaluate training-induced plasticity in the changes in the corticospinal tract, the primary pathway governing voluntary motor control ^38,39^, including PA aftereffects and electrical stimulation^11,40,41^. Other studies have used fMRI^15,42–44^, and EEG^45,46^ to examine neural mechanisms of PA, however, TMS is a convenient and accessible method to compare cortical motor circuits before and after PA. Paired pulse TMS also provides the unique opportunity to evaluate faciliatory and inhibitory intracortical neural circuits that may contribute to overall primary motor cortex (M1) output ^47^.

In the current study, we hypothesized that following a single session of PA, leftward aftereffects of PA will be accompanied by an increase in M1 excitability of left upper limb muscles. Second, we hypothesized that PA-induced behavioral and neurophysiological effects would be enhanced by the addition of somatosensory stimulation to the left upper limb during prism exposure. Finally, we hypothesized that PA would not only induce neurophysiological after-effects in the left upper limb, but the effects of PA will also generalize to the left lower limb. Here, we tested these hypotheses in a group of young neurologically unimpaired individuals, as the first step toward understanding the neural and behavioral mechanism of PA in the neurologically unimpaired nervous system. We used TMS to compare the effects of PA without (Sham) and with somatosensory stimulation (Stim) on overall corticomotor excitability, intracortical circuitry, and visuo-motor behavior.

## 2. Methods

### 2.1 Participants

Sixteen young able-bodied individuals (9 female, 7 male, age 26.7± 4.4 years) participated in a repeated-measures crossover study design involving two conditions: PA combined with sham stimulation (PA+Sham) and with somatosensory electrical stimulation (PA+Stim) (Table 1). The average time between the 2 sessions was 27±8.7 days. One participant withdrew from the study due to discomfort during TMS (Total N= 15). Handedness was determined based on self-report, and 1 participant was left-handed. All participants had normal or corrected-to-normal vision. Participants were excluded if they had a history of or evidence of orthopedic, physical, or neurological pathology, pregnancy (female), presence of skin conditions, bruises, or cuts at the stimulation electrode placement site, impaired sensation in the left upper limb, concurrent enrollment in upper limb rehabilitation or another investigational study, evidence of any medical conditions interfering with study procedures, cardiac pacemaker or other implanted electronic devices, and any contraindications to TMS^40^. All participants provided written informed consent, and the experimental protocol was approved by the Emory University Institutional Review Board and registered on clinicaltrials.gov.

**Table 1:** Study participant demographics.

|  | **Age**<br>**(Years)** | **Gender** | **Race** | **Order of Stim** |  | **Time between**<br>**sessions (Days)** | **Handedness** |
| --- | --- | --- | --- | --- | --- | --- | --- |
|  |  |  |  | **1<sup>st</sup> session** | **2<sup>nd</sup> session** |  |  |
| 1 | 30-35 | Male | White | Sham | Stim | 28 | Right |
| 2 | 20-25 | Female | Asian | Stim | Sham | 16 | Right |
| 3 | 20-25 | Female | White | Stim | Sham | 20 | Right |
| 4 | 20-25 | Female | White | Sham | Stim | 20 | Right |
| 5 | 25-30 | Female | Black | Sham | Stim | 21 | Right |
| 6 | 25-30 | Male | Black | Sham | Stim | 21 | Left |
| 7 | 25-30 | Female | White | Stim | Sham | 29 | Right |
| 8 | 25-30 | Female | Black | Sham | Stim | 18 | Right |
| 9 | 25-30 | Male | Asian | Stim | Sham | 45 | Right |
| 10 | 25-30 | Male | Asian | Sham | Stim | 26 | Right |
| 11 | 20-25 | Male | Asian | Stim | Sham | 35 | Right |
| 12 | 30-35 | Male | Asian | Sham | Stim | 28 | Right |
| 13 | 20-25 | Female | White | Stim | Sham | 33 | Right |
| 14 | 25-30 | Male | Black | Sham | Stim | 19 | Right |
| 15 | 20-25 | Female | White | Stim | Sham | 41 | Right |
|  | 26.7± 4.4 | 7M, 8F | - | - | - | 27±8.7 | - |

### 2.2 Study Procedures

Participants completed two experimental sessions comprising PA+Sham and PA+Stim with approximately a 3-week wash-out period between sessions. We randomized the order of sessions such that half the participants completed PA+Sham first and the other half completed PA+Stim first. (Figure 1. Each experimental session comprised 3 parts in the following order:

1. Pre-PA measurement of cortical neurophysiology using TMS, spatial bias using two visuo-spatial behavioral pointing tasks, with eyes open and the other with eyes closed
2. PA session combined with somatosensory or sham stimulation (PA+Stim or PA+Sham)
3. Post-PA measurement of cortical neurophysiology and visuo-spatial pointing Participants were blinded to when they received Stim or Sham with PA in both sessions.

**Figure 1:**
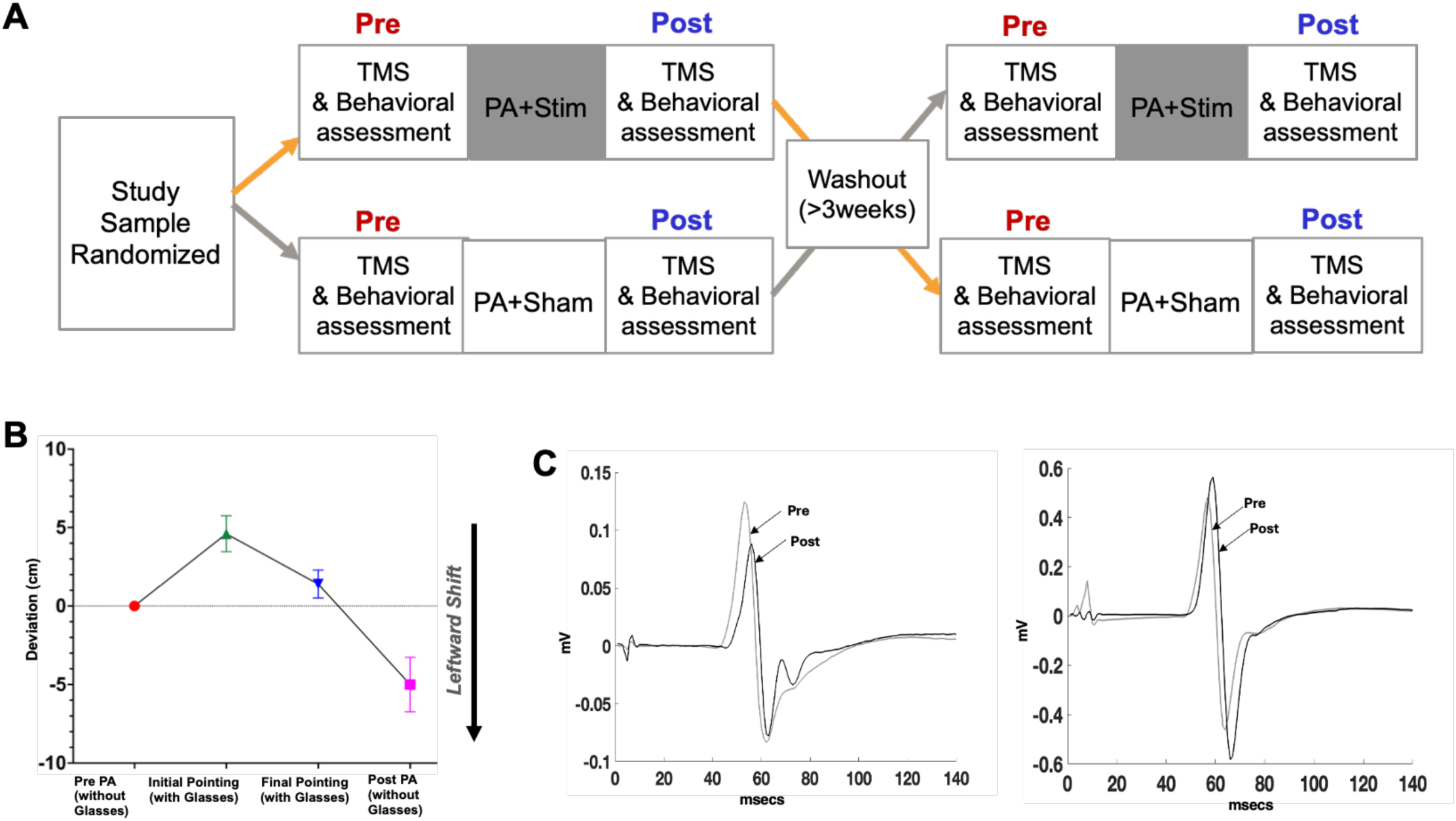
<u>A)</u> Overview of study design: Participants were randomized to receive one session of PA combined with wither Stim (PA+Stim) or Sham (PA+Sham) in a randomized order with at least a 3-week washout period between the two conditions. Behavioral (visuo-proprioceptive and proprioceptive pointing) and neurophysiologic (single and paired-pulse TMS-evoked responses) were obtained Pre and Post each session of PA. **<u>B)</u>** Representative behavioral data during the upper extremity pointing task with eyes open before (without prism glasses on), during (with rightward shifting prism glasses on), and after PA (without prism glasses on). **<u>C)</u>** Representative neurophysiological data showing TMS-evoked MEP responses from the left FDI muscle at pre and post time points in the PA+Sham (left) and PA+Stim (right) conditions.

#### Participant Setup and EMG Placement

Surface electromyography (EMG) signals were recorded from bilateral first dorsal interossei (FDI) muscles (Kendall^©^ 24mm Ag/AgCl, H124sG), left tibialis anterior (TA), and left soleus muscles (Biopac^©^ 35 mm A/AgCl, EL502).

#### Single and Paired Pulse TMS Measurements

During the TMS measurements, participants were in a quiet seated position in a standard chair, back supported legs bent at 90 and hands resting on a firm foam bar while recording from bilateral FDI muscles, and were in a quiet standing position during the recording of left TA and left soleus responses, with both hands resting on a bar. Single monophasic and paired TMS pulses (Magstim 200^2^, MagStim, Wales, UK) were delivered using a 50mm hand-held double circular coil positioned at a 45^0^ angle from the midsagittal plane to target the FDI muscles, and a 70mm figure-of-eight custom batwing coil positioned to target the left TA and left soleus muscles, to induce posterior-anterior (PA) current in M1 in response to TMS. A real-time neuronavigation system (BrainSight, Rogue Research) was used to determine and consistently maintain coil position at the location of bilateral FDI as well as left soleus hotspots, identified as the cortical site on the contralateral motor cortex that generated the largest and most consistent MEPs from the respective targeted muscles. Motor-evoked potentials (MEPs) in response to TMS pulses were collected at the sampling rate of 2000 Hz and band-pass filtered at 10-1000Hz using a 6-channel wireless EMG System (Biopac^©^ Systems Inc. MP160WSW, AcqKnowledge software). The resting motor threshold (RMT) for bilateral FDI and active motor threshold (AMT) for soleus, were determined using the ML-PEST method (maximum likelihood model of parameter estimation by sequential testing (PEST) software^49^.

Resting motor threshold was determined as the minimum intensity that generated ≥ 0.05mA MEP peak-to-peak amplitude for bilateral FDI, and active motor threshold was determined as the minimum intensity that generated >0.1mA MEP amplitude for the TA/soleus. A total of 10 – 20 suprathreshold TMS pulses were delivered to measure corticomotor excitability. Next, three sets of paired TMS pulses were delivered to obtain measures of intracortical circuit excitability-intracortical facilitation (ICF), short intracortical inhibition (SICI), and long intracortical inhibition (LICI). The paired-pulse stimulation parameters were sent at conditioning pulse 80% (FDI) or 90% of motor threshold (soleus and TA), test pulse at 130% above motor threshold, and interstimulus interval 12ms for ICF; conditioning pulse 80% (FDI) or 70% (soleus), test pulse 130%, and interstimulus interval 2ms for SICI; and conditioning pulse 130%; test pulse 130%, and interstimulus interval 100ms for LICI.

Post-PA TMS data were collected using the same hotspot and TMS intensities as Pre-PA. During the second session, we used the TMS hotspot from the initial session, confirmed the hotspot location, and determined the MEP thresholds again to collect the TMS data.

#### Visuo-spatial Motor Behavior Measurements

To assess the sensorimotor effects of PA, the Kessler Foundation Prism Adaptation Treatment (KF-PAT®) protocol and devices were used ^50^. The proprioceptive pointing task requires participants to point straight from the chest with the right upper limb until full arm extension while closing their eyes. The other pointing task, i.e., visuo-proprioceptive pointing, requires participants to point straight from the chest to visual targets placed to their left, right, and center with their arms covered using a cloth to obscure visual feedback of the upper limb and hands.

#### Prism Adaptation

During the prism exposure, participants wore a 20-diopter left-base wedged prism lenses that induced a rightward 12.4° visual field (Bernell™ Deluxe Prism Training Glasses) fitted in goggles that blocked peripheral views, and a visual field occluder blocked the participant’s view of their proximal arm movements. Participants performed 120 pointing movements with their right arm fully extended toward visual targets located at 0^0^ or 21^0^ to the right or left of a board that was oriented to the participant’s median plane.

#### Electrical Stimulation and Sham Stimulation Setup

Surface rectangular electrical stimulation electrode pads (2” x 4”) were placed on the left upper limb over the belly of the triceps and biceps muscles in the upper arm, and smaller 2” x2” square pads were placed on the ventral aspect of the left forearm muscles. Electrical stimulation intensity was customized according to each participant’s sensory threshold, determined as the lowest intensity at which the stimulation was perceived, and motor threshold, determined as the lowest intensity that elicited a visible muscle twitch. During PA+Stim, stimulation was delivered at approximately the midpoint of sensory and motor thresholds for the duration of 20 minutes during prism exposure (i.e. above sensory threshold). During PA+Sham, stimulation electrodes were placed on the arm and forearm as described above, threshold was determined, and then the stimulation device was turned off. Stimulation was delivered using an EmpiÓ TENS unit, at a pulse rate of 100Hz and pulse with of 300μsec.

### 2.3 Data Analyses

Each participant’s neurophysiology data were analyzed using TMS GUI software^42^. Primary behavioral dependent variables included deviation during visuo-proprioceptive pointing and proprioceptive pointing tasks performed with the right upper limb, with deviation toward the left measured as negative numbers and to the right as positive numbers, as well as the distance (in mm) between the line bisection mark and true center of a line. Primary neurophysiologic dependent variables included suprathreshold MEP amplitude for each of the 4 muscles (left FDI, right FDI, left soleus, left TA). Secondary variables included SICI, LICI, and ICF (calculated as the ratio of the conditioned MEP amplitude versus the unconditioned MEP amplitude) for each of the 4 muscles. For the primary variables, we used a 2-way ANOVA to evaluate the effects of 2 levels of training condition (Stim PA, Sham PA) and 2 levels of time (Pre, Post) on each dependent variable, with appropriate Bonferroni-corrected post-hoc comparisons. Additionally, for all variables (primary and secondary), we conducted a priori planned paired t-tests to compare the pre to post change or percentage (%) change scores (Post/Pre x 100) between the PA+Stim and PA+Sham conditions. The significant level (alpha) threshold was set at 0.05 for the statistical tests. We used IBM SPSS^©^ statistics software for statistical analyses (29.0.2.0 (20)).

## 3. Results

### 3.1 Primary Behavioral Outcome Measures: Visuo-proprioceptive and Proprioceptive Pointing

For visuo-proprioceptive pointing, there was a significant main effect of time (F = 100.84, η^2^ = 0.88, p <0.001,), no significant effect of condition (F = 0.43, η^2^ = 0.03, p = 0.524), and no significant condition by time interaction (F = 0.03, η^2^ = 0.00, p = 0.876). The paired t-test comparing pre-post change scores between the PA+Stim and PA+Sham conditions showed no significant difference in the magnitude of training-induced change for PA+Stim (−3.52 ± 6.55 SD) versus PA+Sham (−3.49 ± 7.37 SD) condition (t=-0.056, η^2^ =0.00 p= 0.956,). (Figure 2)

**Figure 2:**
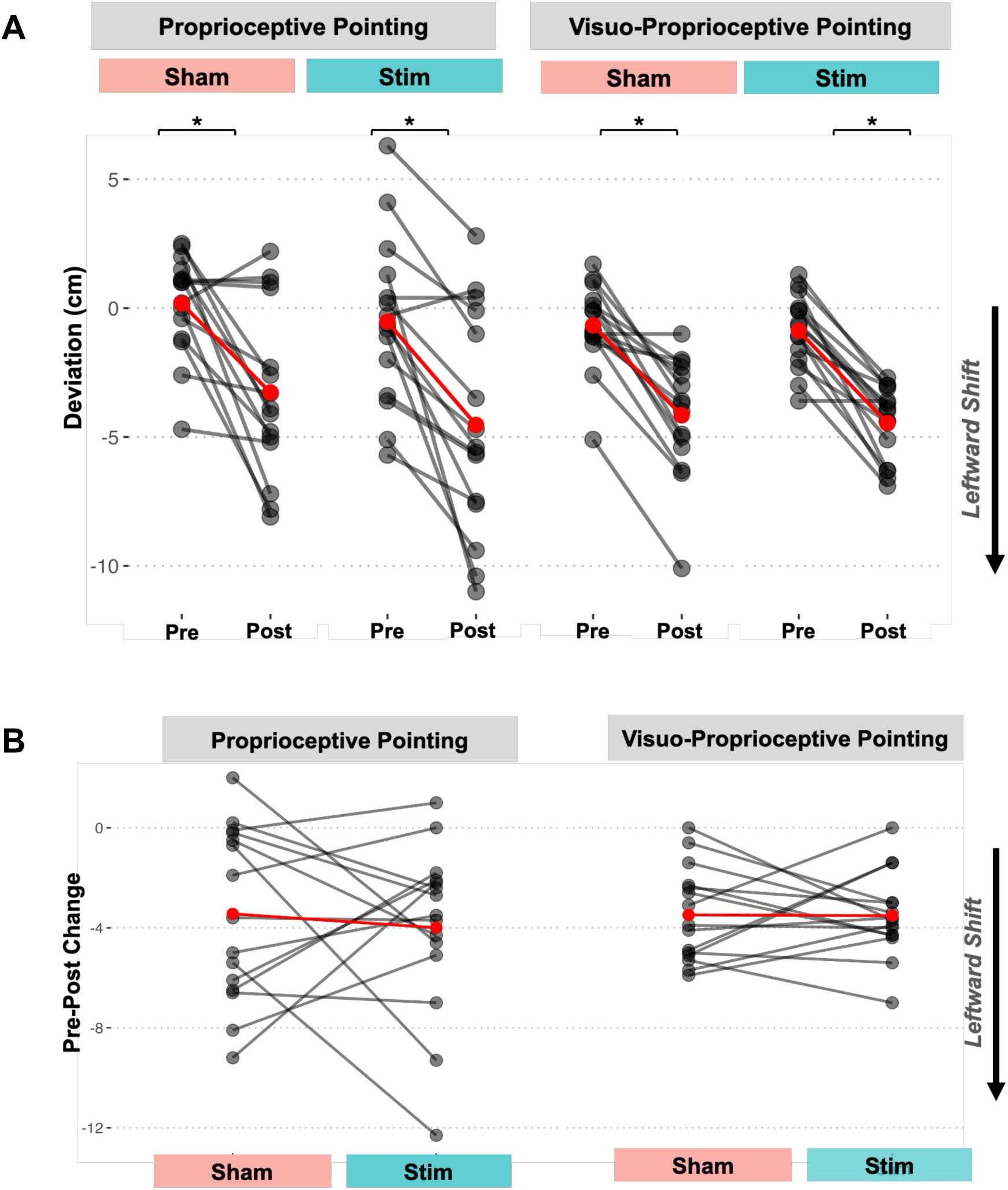
Effects of PA combined with Stim and Sham on visuospatial-motor behavior: **<u>A)</u>**Visuo-proprioceptive pointing (eyes open) performance during the PA+Sham and PA+Stim conditions at pre and post timepoints. Note that there was a significant leftward aftereffect at post versus pre, as shown by a greater negative shift from pre to post for both the PA+Stim condition (p= <0.001) and the PA+Sham condition (p= <0.001). **<u>B)</u>** Proprioceptive pointing (eyes closed) performance during PA+Sham and PA+Stim conditions at the pre and post timepoints. We observed a significant leftward aftereffect, as shown by a shift or deviation toward the left space from pre to post for both the PA+Stim condition (p= <0.001) and for the PA+Sham condition (p= <0.002). **<u>C)</u>** Graph comparing change scores (post minus pre) for visuo-proprioceptive (eyes open) and proprioceptive pointing (eyes closed) for the PA+Sham and PA+Stim conditions. There was no significant difference in magnitude of change between the 2 conditions.

For proprioceptive pointing, there was a significant main effect of time (F= 30.66, η^2^ = 0.69, p = < 0.001), no significant main effect of condition (F= 1.53, η^2^ = 0.1, p= 0.237), and no significant condition by time interaction (F= 0.22, η^2^ = 0.16, p= 0.646). The paired t-test comparing pre-post change scores between the PA+Stim and PA+Sham conditions showed no significant difference in the magnitude of Pre to Post change for PA+Stim (−4.000 ± 13.27 SD) versus the PA+Sham (−3.45 ± 13.49 SD) conditions (t= −0.470, η^2^ =0.02, p= 0.646,) (Figure 2).

### 3.2. Primary Neurophysiological Outcome Measures: TMS-evoked MEPs of bilateral upper limb and left lower limb muscles

For MEP amplitude of the left FDI, there was a significant effect of condition (F = 5.33, η^2^ = 0.29, p= 0.039), no significant main effect of time (F = 0.39, η^2^ = 0.03, p = 0.544), and a significant condition by time interaction (F = 6.33, η^2^ = 0.33, p = 0.026). The post-hoc paired comparisons showed that left FDI MEP amplitude significantly increased from pre (0.875mV ± 0.71 SD) to post (1.655mV ± 1.14 SD) for the PA+Stim condition (η^2^ = 0.52, p=0.002), but showed no significant change from pre (1.101mV ± 1.02 SD) to post (1.083mV ± 0.94 SD) for the PA+Sham condition (η^2^ = 0.00, p= 0.941). The paired t-test comparing pre-post change % scores between the PA+Stim and PA+Sham conditions showed a significantly larger magnitude of change in left FDI MEP amplitudes for the PA+Stim (+0.74mV ± 0.79 SD) vs the PA+Sham (+0.18mV ± 0.56 SD) condition (t = 2.48, η^2^ = 6.88, p = 0.029). (Figure 3)

**Figure 3:**
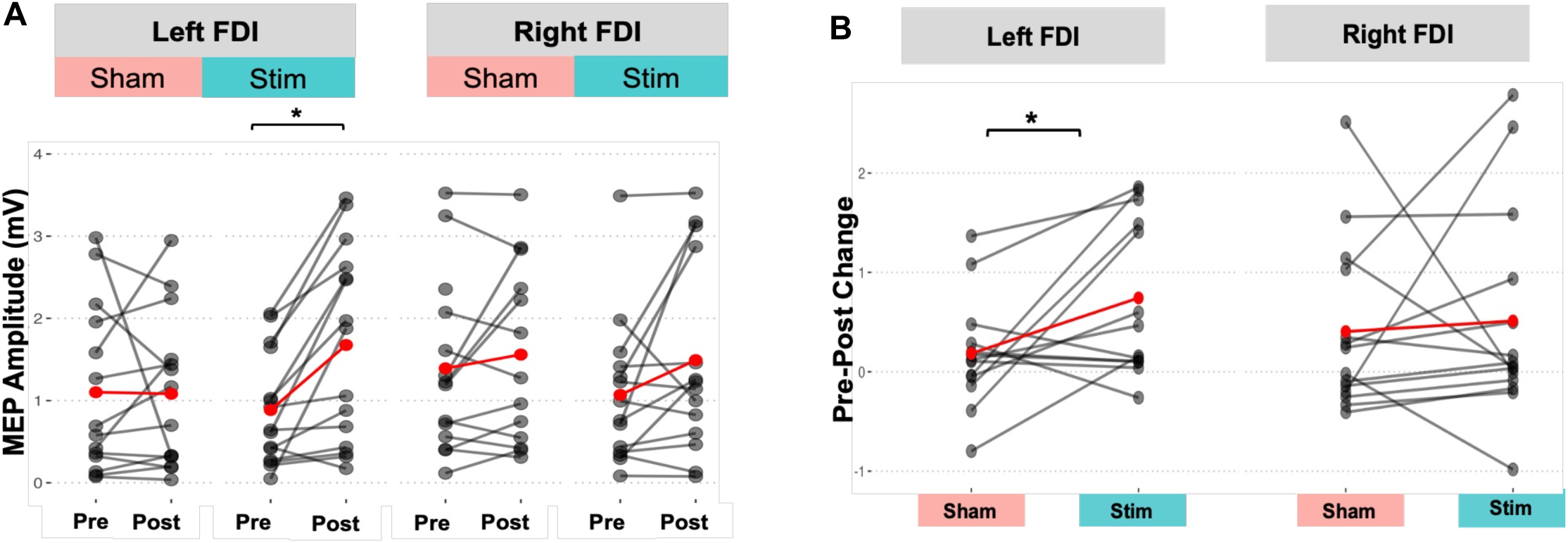
Effects of PA combined with Stim and Sham on corticomotor excitability of upper extremity (FDI) muscles. **<u>A)</u>** MEP amplitudes at Pre and Post timepoints for the PA+Sham and PA+Stim for the left FDI and right FDI muscles. Left FDI MEPs showed a significant increase at Post versus Pre in the PA+Stim condition (p= 0.002). **<u>B)</u>** Training-induced % change scores (post/pre x100) in left FDI and right FDI MEP amplitudes for the PA+Sham and PA+Stim conditions. We found a significantly larger magnitude of change in left FDI MEP amplitudes for the PA+Stim vs PA+Sham condition (p= 0.029, t= 2.482). There was no significant difference between PA+Sham and PA+Stim in the magnitude of change in right FDI MEP amplitudes.

For MEP amplitudes of the right FDI, there was a trend for a significant main effect of time (F= 3.65, η^2^ = 0.23, p= 0.080), no significant effect of condition (F= 0.72, η^2^ = 0.06, p= 0.412), and no significant condition by time interaction (1-β = 0.05, F = 0.004, η^2^ = 0.00, p= 0.951). The paired t-test comparing pre-post % change scores between the PA+Stim and PA+Sham conditions showed that there was no significant difference in the magnitude of change for the PA+Stim (0.31mV ± 0.85 SD) versus PA+Sham (0.41mV ± 0.85 SD) conditions (t = −0.30, η^2^ = −0.08, p = 0.768). (Figure 3)

For left TA MEP amplitudes, there was no significant main effect of time (F =2.74, η^2^ = 0.2, p= 0.126), no significant main effect of condition (F= 1.52, η^2^ = 0.12, p = 0.244), and no significant condition by time interaction (1-β = 0.05, F = 0.001, η^2^ = 0.00, p= 0.98). The paired t-test comparing pre-post % change scores showed no significant difference in the magnitude of Pre to Post change in TA MEP amplitudes for the PA+Stim (0.11mV ± 0.16 SD) versus PA+Sham (0.11mV ± 0.46 SD) conditions (t = −0.026), η^2^ = 0.00, p = 0.98). (Figure 4)

**Figure 4:**
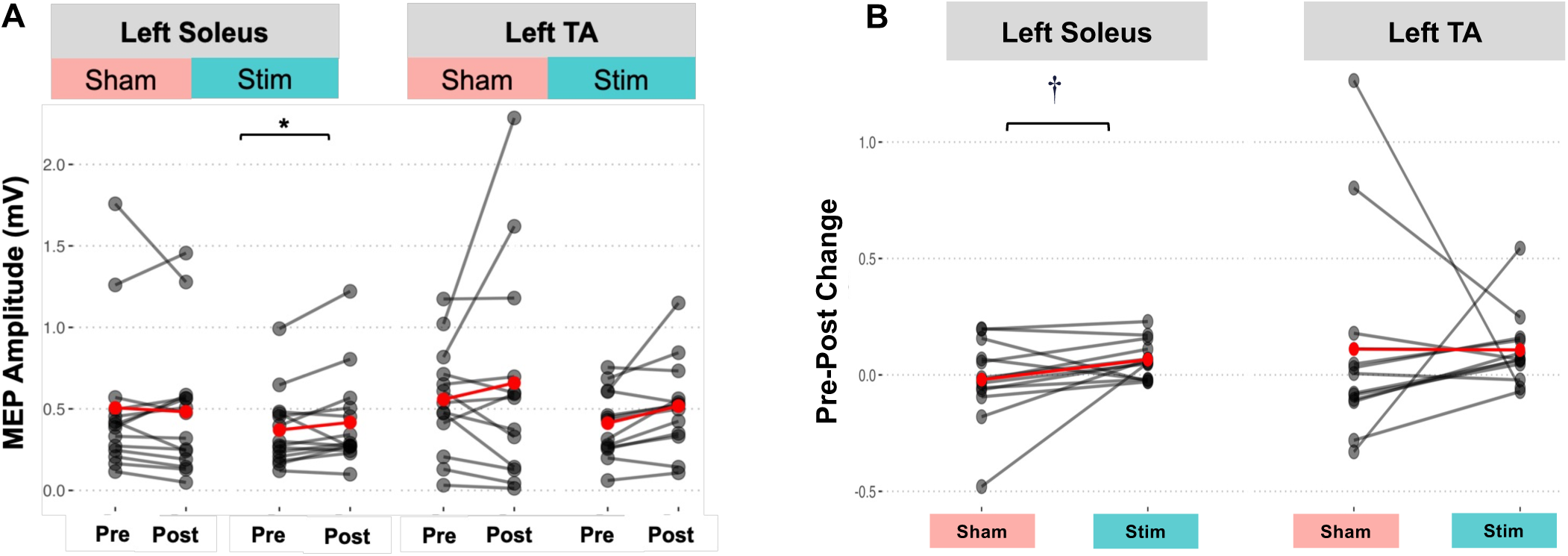
Effects of PA combined with Stim and Sham on corticomotor excitability of left lower extremity (soleus, TA) muscles. **<u>A)</u>** MEP amplitudes at Pre and Post timepoints for the PA+Sham and PA+Stim for the left Soleus and left TA muscles. Left soleus MEPs showed a significant increase for the PA+Stim condition (p= 0.014). There was no significant difference in left TA MEPs between the 2 conditions.**<u>B)</u>:** Training-induced % change scores (post/pre x100) in left soleus and TA MEP amplitudes for the PA+Sham and PA+Stim conditions. Pre-post % change scores for left soleus MEP amplitude showed a statistical trend for a larger magnitude of change for PA+ Stim (p= 0.093, t=1.823). There was no significant difference between PA+Sham and PA+Stim in the magnitude of change in left TA MEP amplitudes.

For left soleus MEP amplitudes, there was no significant main effect of time (F = 0.55, η^2^ = 0.04, p= 0.472,), no significant main effect of condition (F = 2.17, η^2^ = 0.15, p = 0.166), there was a statistical trend for a condition by time interaction (1-β = 0.39, F = 3.32, η^2^ = 0.22, p= 0.093). The post-hoc paired comparisons showed that left soleus MEP amplitude significantly increased from pre (0.36mV ± 0.25 SD) to post (0.43mV ± 0.32 SD) for the PA+Stim condition (η^2^ = 0.41, p = 0.014), but showed no significant change from pre (0.54mV ± 0.49 SD) to post (0.52mV ± 0.45 SD) for the PA+Sham condition ((η^2^ = 0.01, p= 0.688). (Figure 4)

### 3.3. Secondary Neurophysiologic Variables: ICF, SICI, and LICI

For ICF of the right FDI, the paired t-test comparing Pre to Post % change between the PA+Stim and PA+Sham conditions showed a statistical trend for larger magnitude of change for the PA+Stim (+13.76mV ± 56.47 SD) versus the PA+Sham (−17.49mV ± 33.76 SD) condition (t = 1.80, η^2^ = 0.48, p = 0.095). However, for ICF of the left TA, the paired t-test showed a trend for larger magnitude of change for PA+Sham (39.83mV ± 76.46SD) vs PA+Stim (1.67mV ± 24.99 SD) conditions (t = −1.75, η^2^ = −051, p = 0.11). For ICF of the left FDI and left soleus, the paired t-test comparing Pre to Post % change showed no significant difference between the PA+Stim and PA+Sham conditions (all p-values >0.1). (Figure 5)

**Figure 5:**
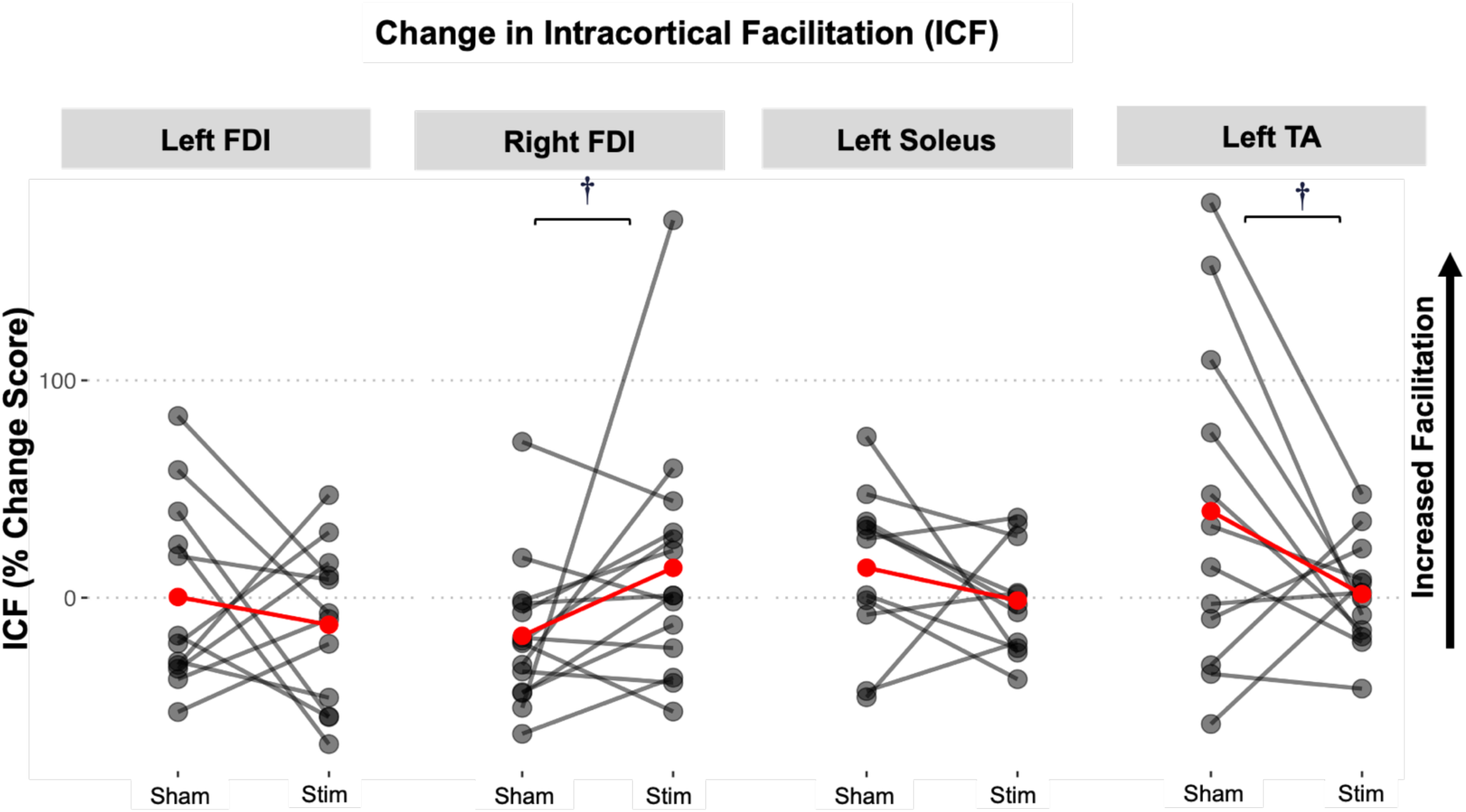
Effects of PA combined with Stim and Sham on intracortical facilitation (ICF). The figure shows graphs showing training-induced change scores (post/pre X100) ICF for the PA+Sham and PA+Stim conditions for Left FDI **<u>(A),</u>** right FDI **<u>(B),</u>** left TA **<u>(C),</u>** and left soleus**<u>(D)</u>** muscles. Pre-post % change scores for right FDI ICF showed a statistical trend for a larger magnitude of change for PA+ Stim (p= 0.095, t=1.80). However, pre-post % change scores for left TA ICF showed a statistical trend for a larger magnitude of change for PA+ Sham (p= 0.11, t=-1.75). There was no significant difference between PA+Sham and PA+Stim in the pre-post % change score in left FDI and left soleus.

For SICI in the left TA, the paired t-test comparing Pre to Post % change showed a significant larger magnitude of change for the PA+Sham (+21.36mV ± 36.92SD) vs PA+Stim (−29.18mV ± 26.01 SD) condition (t = −3.24, η^2^ = −1.03, p = 0.01). For the left FDI, right FDI, and left soleus muscles, the paired t-test comparing Pre to Post % change in SICI showed no significant differences between the PA+Stim and PA+Sham conditions (all p-values >0.1). (Figure 6)

**Figure 6:**
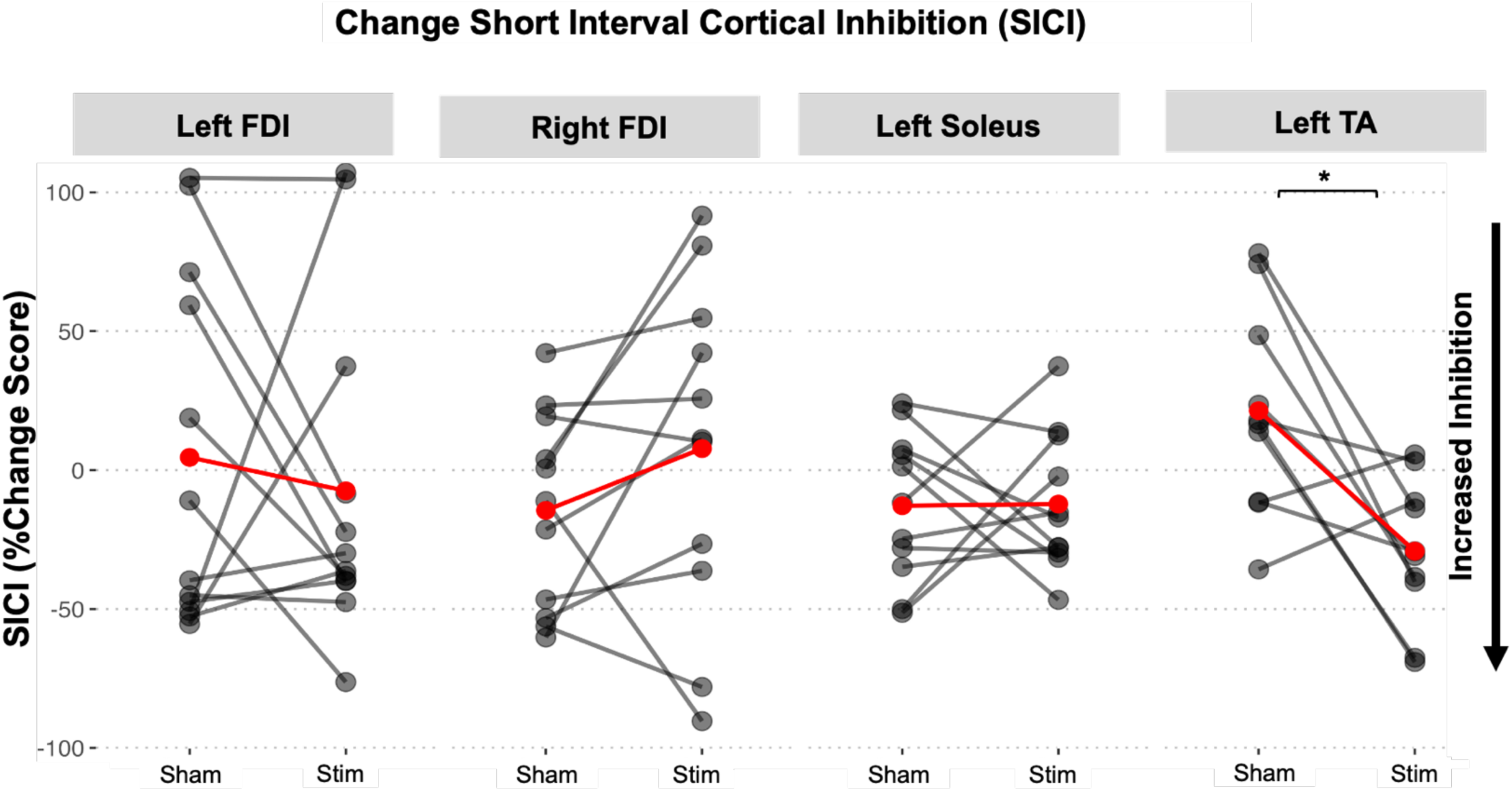
Effects of PA combined with Stim and Sham on short interval intracortical facilitation (SICI). The figure shows graphs showing training-induced change scores (post/pre X100) SICI for the PA+Sham and PA+Stim conditions for Left FDI **<u>(A),</u>** right FDI **<u>(B),</u>**left TA **<u>(C),</u>** and left soleus **<u>(D)</u>** muscles. Only pre-post % change scores for the left TA showed a larger magnitude of change for PA+ Sham (p= 0.01, t=-3.24).

For LICI in the left FDI, the paired t-test comparing Pre to Post % change scores showed a significant larger magnitude of change for the PA+Sham (+91.29mV ± 179.62 SD) vs PA+Stim (−26.87mV ± 56.47 SD) condition (t = −2.26, η^2^ = −0.65, p = 0.045). For LICI of the left TA, the paired t-test comparing % change scores showed a trend for a larger magnitude of change for the PA+Sham (+26.28mV ± 78.73 SD) versus PA+Stim (−24.16mV ± 27.05 SD) condition (t = −1.74, η^2^ = −0.55, p = 0.116). For the right FDI and left soleus muscles, the paired t-test comparing Pre to Post % change in LICI showed no significant difference between the PA+Stim and PA+Sham conditions (all p-values >0.1). (Figure 7).

**Figure 7:**
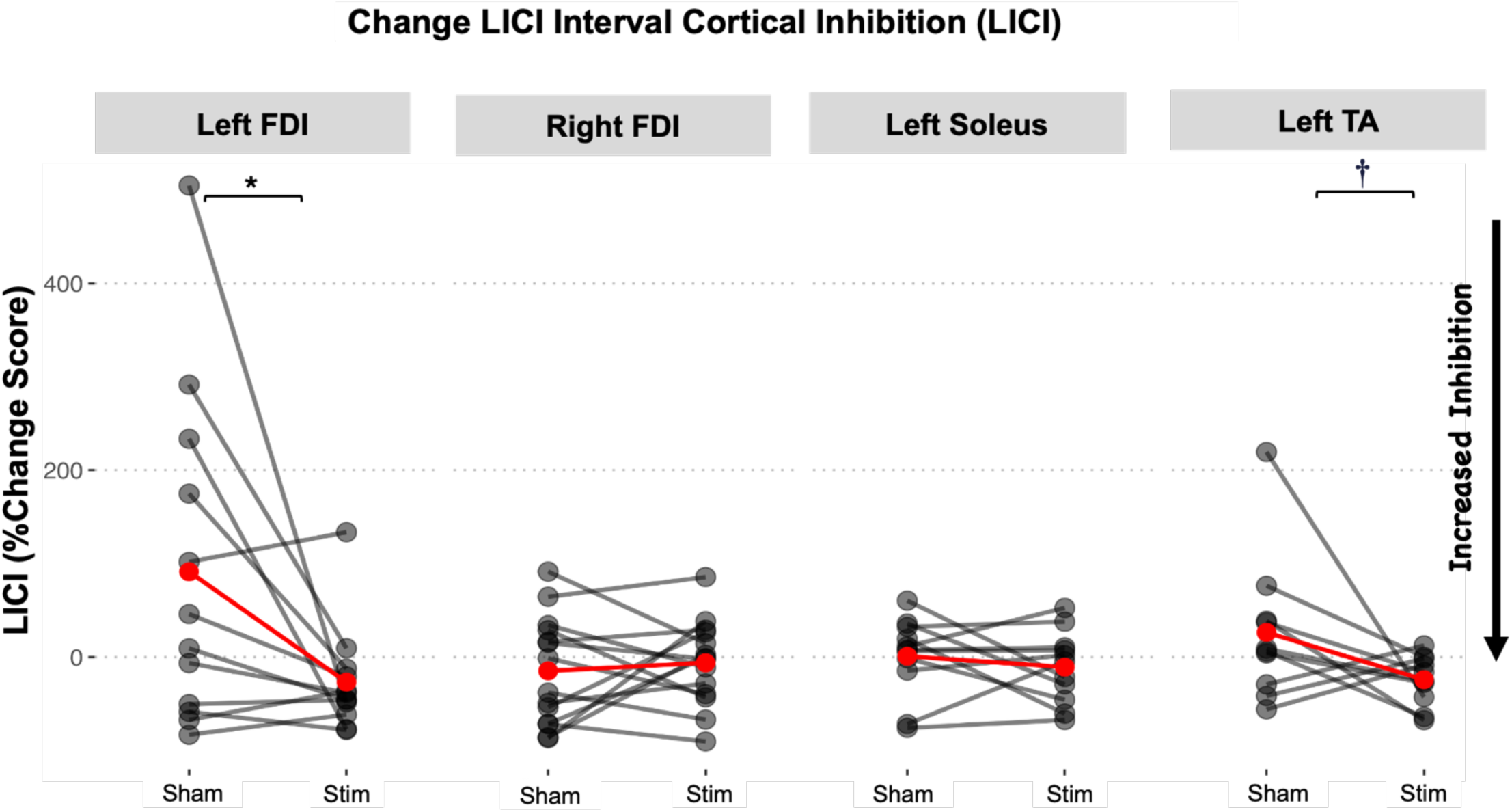
Effects of PA combined with Stim and Sham on long interval cortical inhibition (LICI). The figure shows graphs showing training-induced change scores (post/pre X100) for the PA+Sham and PA+Stim in the left FDI **<u>(A),</u>** right FDI **<u>(B),</u>** left TA **<u>(C),</u>** and left soleus **<u>(D)</u>.** Pre-post % change scores for left FDI LICI showed a larger magnitude of change for PA+ Sham (p= 0.045, t=-2.26). Similarly, pre-post % change scores for left TA SICI showed a statistical trend for a larger magnitude of change for PA+ Sham

## 4. Discussion

We evaluated the behavioral and neural effects of a single session of PA with right-shifting prism glasses (to induce leftward aftereffects) in combination with somatosensory electrical stimulation (PA+Stim) or sham stimulation (PA+Sham) delivered to the left upper limb (toward the direction of the PA aftereffect). Our results showed that in young healthy adults, as expected, a single session of both PA+Stim and PA+Sham induced a significant visuo-motor behavioral aftereffect, measured as a leftward shift during visuo-proprioceptive and proprioceptive pointing tasks. We observed no differences between PA+Stim versus PA+Sham conditions in these behavioral after-effects. Our neurophysiologic data showed that the PA+Stim condition increased MEP amplitudes of the left FDI (toward the direction of the prismatic after-effect), while the PA+Sham condition did not induce significant changes in left FDI MEP amplitudes. We also showed that only the PA+Stim condition increased MEP amplitudes in the left soleus muscles, with no significant change observed for the PA+Sham condition. Furthermore, the PA+Stim condition induced a larger increase in intracortical facilitation (measured via ICF) in the left FDI with a trend for a larger change for the left tibialis anterior. PA+Sham induced a significantly larger magnitude of training-induced change in SICI for the left tibialis anterior, LICI for the left FDI, and a statistical trend for a larger change in SICI for the left tibialis anterior.

### Behavioral aftereffects of PA were observed for both the Sham and Stim conditions

As expected, our results showed that right-shifting PA (both with Stim and Sham) induced a leftward aftereffect leading to a leftward shift in visuo-spatial orientation during upper limb pointing movement tasks in young healthy adults. This finding is consistent with our hypothesis and previous literature, supports the robust behavioral effects induced by only one session of PA, and shows the promise of PA as an intervention paradigm targeting visuo-spatial orientation and motor function^6,11,53^. We observed similar behavioral effects following PA+Stim and PA+Sham, suggesting that adding somatosensory stimulation to PA may not induce marked augmentation of PA’s behavioral effects within only one session.

### PA induced significant changes in corticomotor excitability of the Left and Right FDI

We had hypothesized that PA will increase corticomotor excitability of the left FDI (toward the direction of prismatic aftereffect). Our prediction was based on the rationale that PA biases visuospatial motor behavior toward the left hemi-space by inducing changes in the right dorsal fronto-parietal networks (within the superior parietal lobule/intra-parietal sulcus and the dorsolateral prefrontal cortex) responsible for visuospatial attention and orientation, which in turn have connectivity to the primary motor cortex^48^. Our results supported our hypothesis, but only for the PA+Stim condition, as shown by a significant increase in contralateral corticomotor excitability after PA+Stim, and a significantly larger training-induced change in left FDI MEP amplitudes with PA+Stim versus PA+Sham. Our results are somewhat consistent with another study^11^ that evaluated the behavioral and neural mechanisms of PA (alone) using TMS, because they also found that although the rightward shifting (leftward aftereffect) PA had behavioral aftereffects, PA did not significantly increase corticomotor excitability in the left FDI in young healthy adults.^11^

Previous theoretical models of intra- and inter-hemispheric interaction in people with neglect^54^ suggest that right hemispheric visual attentional damage causes a functional imbalance within the left and right dorsal parieto-frontal attentional networks, with decreased activity in the right dorsal parieto-frontal network and subsequent hyperactivity of the left dorsal parieto-frontal network, creating a right-sided attentional circuit bias and left-sided neglect. Therefore, we hypothesized that following PA, there would also be a decrease in ipsilateral corticomotor excitability (i.e. right FDI MEP amplitude would decrease) in addition to an increase in left FDI corticomotor excitability. Our results did not support this hypothesis, as our results in fact showed an increase no change in right FDI corticomotor excitability following PA+Sham. The findings of Magnani et al^11^ were different from our current results, as they showed that the leftward aftereffect induced by PA was not accompanied by any significant change in corticomotor excitability in the ipsilateral (right) FDI.

### Neurophysiological aftereffects of PA combined with upper limb motor training and Stim may carry over to the left lower limb

As stated above, we predicted that as rightward shifting PA will increase excitability in the left visuo-spatial attentional networks, the resulting enhancement in M1 output could potentially be generalized or carried over to the left lower limb, and not just be localized to the left upper limb. Thus, we evaluated whether PA resulted in increased corticomotor excitability for both left upper and left lower muscles, which, to our knowledge, has not been evaluated before. Somewhat consistent with our hypothesis, we did observe significantly increased corticomotor excitability in the left TA and left soleus muscle for the PA+Stim condition. Our results suggest that when PA is combined with somatosensory stimulation, we may see a greater magnitude of carryover of PA to lower limb neural output. Potentially, combining PA with a lower limb motor training task or adding somatosensory stimulation to the left lower limb may further augment these carryover effects to lower limb muscles, and merit more investigation. Our results suggest that more research is needed to evaluate and target lower limb neurophysiology following the administration of PA in individuals with post-stroke neglect. Additionally, because behavioral changes may accompany corticomotor excitability, we may also need to examine the corresponding effects of PA on lower muscle performance tasks such as leg pointing or placement with walking.

### Neurophysiological aftereffects of PA on intracortical excitability and inhibition

We found no changes in ICF in bilateral upper limb or left lower limb muscles following either PA+Sham or PA+Stim. Notably, there was high inter-individual variability in the paired-pulse ICF data for both upper limbs and left upper limb, which may have influenced these results. Notably, however, SICI of the left TA increased significantly, and SICI of left soleus showed a statistical trend toward an increase in PA+Stim training condition. Previously, SICI has been shown to change during different brain states, increasing during rest, and reducing just before voluntary contraction or simple reaction task^55,56^ in able-bodied individuals. Potentially, PA-induced increases in SICI may be partly related to the TMS measures being collected at rest. However, in contrast to our findings, Magnani et al^11^ showed that rightward shifting PA alone (leftward aftereffect) produced an increase in ICF and no changes in SICI of the left upper limb muscles. Our study did not show any PA-induced changes in LICI. No previous studies have investigated the effects of PA on LICI. The effects of PA combined with Stim on intracortical inhibitory and facilitatory circuits merits more investigation in larger sample studies.

### Adding Stim as an adjuvant to PA may augment PA-induced neuroplasticity and enhance generalization to lower limb

Previously, sensorimotor electrical stimulation has been shown to increase afferent input in the somatosensory cortex^57,58^, which in turn can enhance the excitability of motor (M1)^34–36^, and possibly visuo-spatial^28,29^ cortical areas. Thus, we predicted that PA+Stim will further enhance the upregulatory effects of PA on left upper limb M1 (because stim is being delivered to the (left) UE), leading to more robust neurophysiological and behavioral after-effects with PA+Stim. Consistent with our hypothesis, we showed that PA+Stim indeed increased left FDI MEP amplitude. Furthermore, PA+Stim resulted in a significantly larger training-induced change in MEP amplitude of the left FDI compared to PA+Sham (Figure 1, Panel B). Our current results may be somewhat in congruence with other studies which showed that PA combined with left neck extensor muscle somatosensory feedback (neck vibration stimulation)^59^ as well as left^60^ and right^61^ M1 anodal cortical stimulation (a-tDCS) may have more robust effects on spatial neglect deficits and motor function than PA alone in chronic post-stroke individuals with spatial neglect ^54^. Somewhat surprisingly, we did not observe corresponding larger behavioral aftereffects of PA+Stim compared to PA+Sham. Potentially, the greater increase in corticomotor excitability induced by PA+Stim versus PA+Sham may require more dosage (e.g. 5-10 sessions) to induce a behavioral change. Thus, we propose that future work should investigate the neural and behavioral additive effects of PA+Stim.

Furthermore, our current findings of significant effects of PA+Stim on corticomotor excitability of leg muscles suggest that right shifting PA (with leftward aftereffect) in conjunction with somatosensory stimulation has carryover effects to the corticomotor excitability and SICI of the left leg. We found that the MEP amplitudes of both the left TA and left soleus significantly increased from pre to post for the PA+Stim condition but not for the PA+Sham condition (Figure 2). Thus, potentially, when somatosensory stimulation is added to PA, the stimulation-induced changes in neural circuit excitability augment the PA-induced neuroplasticity, increasing the likelihood of carryover to lower limb muscles. Interestingly, left soleus and left TA SICI also showed significant decreases after PA+Stim, indicating greater intra-cortical inhibitory influences on M1 after PA+Stim. While another study showed that somatosensory stimulation to the median nerve (wrist) alone induced no changes in SICI or ICF^62^; to our knowledge, no other studies have looked at the effect of PA+Stim on both upper and lower limb muscles.

### Clinical and research implications

While our behavioral data show that both PA+Stim and PA+Sham conditions induced similar sensorimotor aftereffects, the neurophysiological data show that the PA+Stim induced an increase in corticomotor excitability in the left FDI, left TA, and left soleus. The larger neurophysiological effects of PA+Stim suggest that this combined training paradigm might induce a more robust behavioral aftereffect that improves right hemispheric corticomotor excitability, perhaps with multiple PA+Stim sessions. PA+Stim potential to enhance corticomotor connections in the left space with transfer to the left lower limb presents a promising treatment avenue for patients with spatial neglect with motor deficits.

### Limitations

The same order was used for neurophysiological and behavioral tests for all participants. Possibly by the time the last post-test was done (neurophysiology testing), some of the acute effects of PA could have been washed out or weakened, especially in young healthy adults due to the unimpaired neurophysiological system affecting the longevity and robustness of PA effects. However, our concerns were somewhat allayed by previous studies suggesting that the effects of 1-2 sessions of PA last from 24 hours to up to 1 week in post-stroke spatial neglect individuals. Because corticomotor excitability and inhibition can be state-dependent, our SICI, LICI, and ICF results collected at rest (left and right FDI) and quiet standing (TA/soleus) may be different compared to similar measures collected in an active state^63^. In our study, somatosensory stimulation was delivered to the left upper limb at supra-sensory but sub-motor threshold. Future studies can evaluate the effects of stimulation on PA with stimulation delivered at stronger intensities above motor threshold. Similarly, future work can evaluate the effects of combining PA with stimulation delivered to different muscles such as neck or leg muscles in addition to or instead of the upper limb.

Consistent with previous PA literature, the behavioral training task done during PA was performed only with the right arm to match our PA protocol to the protocols used in stroke participants t with left spatial neglect, whose left arm may be paretic. However, this motor training paradigm may not be the most optimal as the left arm does not engage in motor practice, potentially reducing the neural and/or behavioral effects induced by PA. Additionally, the average session duration ranged from three to four hours, so fatigue could have affected the post-test behavioral and neurophysiologic data in some participants. Lastly, the lack of significant cognitive aftereffects of PA may be due to the fact that the participants had an intact neurophysiological system, and future studies should explore the cognitive aftereffects of PA in post-stoke individuals with spatial neglect.

### Conclusions

Using a dose-matched, repeated-measures, sham-controlled study design, our results suggest that PA+Stim may yield larger neurophysiological effects compared to PA alone. Our study provided new evidence that PA may increase corticomotor excitability of the left upper limb (toward the direction of the leftward aftereffect) in young healthy adults, suggesting that PA modulated connections between visuospatial cortical circuits and the primary motor cortex. This study also revealed that neural effects of PA may transfer from the left upper limb to the left lower limb in young healthy adults, especially when PA is combined with Stim. These carryover effects of PA to lower limb have both clinical and neurophysiological implications on the potential uses of PA in the rehabilitation of lower limb and gait deficits in individuals with post-stroke spatial neglect. Future studies could evaluate whether PA+Stim enhances the effectiveness of PA in individuals with spatial neglect, especially in people who do not respond to traditional PA alone. Additionally, based on our current results, the effects of combining PA with Stim on carryover effects to lower limb muscles should be further investigated and may inform the design of novel rehabilitative strategies to improve lower limb and walking function. Future research is needed to elucidate the mechanisms of PA and to optimize the therapeutic protocols of PA and adjuvant stim or motor training for clinical applications.

## Data Availability

All data produced in the present study are available upon reasonable request to the authors

